# ELAVL1: A Novel Diagnostic and Prognostic Biomarker in Lung Adenocarcinoma Linked to Ferroptosis

**DOI:** 10.1101/2025.11.23.25340837

**Authors:** Xiaohai Li, Yi Hu, Bo Shao, Jian Wang

**Author notes:** Corresponding author Xiaohai Li: Department of Thoracic Surgery, Zhaoqing First People’s Hospital, NO.9 Donggang East Road, Zhaoqing 526000, China. Jian Wang: Department of Thoracic Surgery, Affiliated Cancer Hospital and institute of Guangzhou Medical University, NO.78 Heng Zhigang Road, Guangzhou 510060, China.

## Abstract

**Background:** Lung adenocarcinoma (LUAD) is the most common malignant neoplasm with a poor 5-year survival rate (<16%). Ferroptosis, an iron-dependent cell death mechanism, has emerged as a key regulator of tumor progression, yet its clinical relevance in LUAD remains unclear. This study aimed to explore ferroptosis-related genes (FRGs) as diagnostic and prognostic biomarkers for LUAD.

**Methods:** Differentially expressed genes (DEGs) were identified from TCGA (526 LUAD vs. 59 normal) and GEO (GSE31210, 226 LUAD) datasets. Hub FRGs were selected via intersection of DEGs and FerrDb V2 genes, followed by survival analysis. Diagnostic models were built using SVM with ten-fold cross-validation, while prognostic signatures were developed through LASSO Cox regression. Immune infiltration (CIBERSORT) and functional enrichment (GO/KEGG) analyses were performed. ELAVL1 and SLC7A11 were validated by IHC in tissues and siRNA knockdown in PC9 cells.

**Results:** The two-gene diagnostic model (ELAVL1 + SLC7A11) achieved high accuracy (AUC = 0.9675). High-risk patients (elevated ELAVL1/SLC7A11) showed poorer overall survival (median OS: 32.1 vs. 48.6 months, HR = 1.78, P < 0.001). Functional enrichment revealed cell cycle dysregulation and immune activation in high-risk groups. ELAVL1 knockdown suppressed LUAD cell proliferation/migration and downregulated CDK4/6.

**Conclusion:** ELAVL1 serves as a dual-functional biomarker for LUAD diagnosis and prognosis, mechanistically linking ferroptosis to immune evasion and cell cycle progression. The two-gene signature provides a robust tool for clinical risk stratification, highlighting ELAVL1 as a potential therapeutic target.

## Introduction

Lung adenocarcinoma (LUAD), the most prevalent form of non-small cell lung cancer (NSCLC), represents nearly 80% of lung cancer cases and has the highest incidence and mortality rates.^1,2^ Despite treatments such as chemotherapy, radiotherapy, and targeted therapy, the 5-year overall survival rate for LUAD patients remains around 16%.^3^ Therefore, understanding the molecular mechanism of LUAD and identifying key target molecules improves the ability to predict patient prognosis.

Ferroptosis, an iron-catalyzed form of regulated cell death, results from disrupted cellular redox balance,^4^ leading to lipid peroxidation and accumulation of iron-dependent lipid hydroperoxides,^5^ which eventually cause cytological changes.^6^ Recent studies have linked ferroptosis to the pathophysiology of diseases like cancer, neurological disorders, ischemia-reperfusion injury, and renal damage.^7–10^ Moreover, compelling evidence indicates that ferroptosis exerts a vital function in suppressing tumors and managing metastasis, thereby holding substantial potential for cancer treatment and prognosis prediction.^11, 12^ Ferroptosis exerts a regulatory impact on the advancement of various tumor types, including renal cancer, triple-negative breast cancer, non-small cell lung cancer, and diffuse large B cell lymphoma.^13–16^ Consequently, inducing ferroptosis has emerged as a viable therapeutic approach against cancer. Therefore, this process may serve as a novel target in the treatment of LUAD.

In this study, hub genes were identified through the intersection of differentially expressed genes (DEGs) and ferroptosis-related genes, followed by survival analysis. Subsequently, we constructed a diagnostic model based on the RNA-seq expression profile of 594 samples from TCGA database. A ferroptosis-related prognostic signature for the prognosis of LUAD patients was further established through univariate and multivariable Cox regression analyses. Moreover, we investigated the characteristics of the gene signature in the tumor microenvironment through functional and immune infiltration analysis. Finally, we assessed the expression of hub genes in clinical tissue samples and explored the possible roles and functions of core genes in LUAD.

## Methods

### Ethics Approval and Consent to Participate

All participants provided informed consent for inclusion before participating in the study, which was conducted in accordance with the Declaration of Helsinki, and the protocol was approved by the Ethics Committee of the Zhaoqing First People’s Hospital (No. 2021-06-06)

### Data Download and Preprocessing

The RNA-seq count data of 585 samples, comprising 526 LUAD and 59 normal lung samples, were downloaded from the Cancer Genome Atlas (TCGA) database (https://portal.gdc.cancer.gov/). The data were then normalized to transcripts per kilobase million value using the Caret package in R (version 4.2.2). Clinical information for LUAD samples was obtained from cBioPortal (https://www.cbioportal.org/), and preprocessing involved removing samples with missing survival state and follow-up information for overall survival time (OS). And a total of 154 genes associated with ferroptosis were retrieved from the FerrDb V2 database (http://www.zhounan.org/).

### Identification of Differentially Expressed Ferroptosis-Related Genes (DEFGs)

The “DESeq2” package was used to analyze the differentially expressed genes (DEGs) between LUAD and normal samples. The threshold was set as |log_2_(FoldChange)| ≥ 2 and adjusted *P*-values < 0.05. Visualization of the DEGs was accomplished with the ‘ggrepe1’ and ‘ggplot2’ packages. The DEFGs, obtained from intersecting DEGs and ferroptosis-related genes, were visualized through the ‘pheatmap’ and ‘ggplot2’ packages. An unpaired Wilcoxon-test was conducted to compare DEFG expression in normal and cancerous tissues using RNA sequencing data in TPM format from the TCGA database.

### Survival Analysis of the DEFGs

Tumor samples from the TCGA database were used with the ‘survival’ R package to study the roles of the DEFGs in OS. Patients were categorized into high- and low-expression groups based on the median expression level of the DEFGs. Kaplan-Meier (KM) survival curves were plotted to compare OS between the two groups, and the log-rank test was used to evaluate statistical significance.

### Building and Testing the Diagnostic Model

A diagnostic model was developed using a support vector machine (SVM) to differentiate LUAD tumors from normal samples, utilizing a two-gene signature. The SVM model was established using the scikit-learn package in Python and trained through ten-fold cross-validation.^17, 18^ The classification prediction’s diagnostic capability was assessed by examining the area under receiver operating characteristic (ROC) curve for both the training and testing datasets from the TCGA database.

### Development and Verification of the Prognostic Model

LUAD patients with OS from TCGA database were included in the clinical correlation analysis. Using the R package ‘caret’, LUAD patients, the total set, were randomly split into training and testing sets in a 1:1 ratio. A prognostic signature was developed using the training set based on the expression levels of SLC7A11 and ELAVL1, with the testing and total sets used for internal validation. The formula for calculating the risk score to predict the prognosis of LUAD patients was established: risk score = ∑β*_i_* × exp_gene*_i_*, the expression level of the gene is represented by exp_gene*_i_*, with β*_i_* as its coefficient. Based on the median of risk score, LUAD patients from the training, the testing and total sets were stratified into high- and low-risk groups. The R package ‘survminer’ was used to conduct survival analysis and plot KM survival curves for the high- and low-risk groups. We evaluated the feasibility and precision of our prognostic model and clinical features in forecasting patient outcomes by calculating the area under the curve (AUC) values with the R package ‘survivalROC’. Ultimately, we carried out univariate and multivariate Cox regression analyses, incorporating clinical information and the risk score.

### Functional Analysis

The ‘ClusterProfiler’ R package was utilized for functional annotation of Gene Ontology (GO) and also for conducting Kyoto Encyclopedia of Genes and Genomes (KEGG) analysis,^19^ a false discovery rate (FDR) ≤ 0.05 were considered statistically significant. Subsequently, LUAD samples were divided into high-risk and low-risk categories according to their risk scores obtained from the TCGA database. Gene Set Enrichment Analysis (GSEA) was conducted to determine the KEGG pathways in these groups. The significance of the pathway was established with *P* < 0.05, normalized enrichment scores (NES) > 1.0, and FDR < 0.25.

### Estimation of Infiltrating Immune Cells

The CIBERSORT algorithm was used to estimate tumor-infiltrating immune cells (TIICs) in both high-and low-risk groups.^20^ The algorithm used normalized gene expression data along with the annotated gene signature matrix (LM22) (https://cibersort.stanford.edu/) to identify 22 immune cell subtypes across various samples. The TIICs were considered significant between the two groups at *P* < 0.05.

### Immunohistochemical Staining Analysis

LUAD and corresponding normal lung tissues from patients were preserved in 10% formalin. Then, 3 μm-thick sections were created using standard procedures and stained with an immunohistochemistry kit (MX Biotechnologies, Fuzhou, China). The tissue sections were incubated overnight at 4 ℃ with rabbit antibodies against SLC7A11 (1:200, HuaBio, Wuhan, China) and ELAVL1 (1:200, HuaBio). The immunostaining score was assessed by two pulmonary pathologists under a blind protocol. For each specimen, the overall score is calculated by multiplying the intensity expression score (0 for no staining, 1 for weak staining, 2 for moderate staining, and 3 for strong staining) by the number of stained cells (1 for ‘≤ 25% of cells,’ 2 for ‘26–50% of cells,’ 3 for ‘51–75% of cells’). A total score >6 points was defined as high expression, otherwise, it was categorized as low expression.^21^

### Real-Time Quantitative Polymerase Chain Reaction (RT-qPCR)

Cell samples were used to isolate total RNA with an RNA isolation reagent from Sigma-Aldrich, Abingdon, MD, USA. Then, 1 µg of RNA was converted into cDNA using a cDNA synthesis kit (Takara, Dalian, China). RT-qPCR was carried out with SYBR Green qPCR Master Mix (Roche, Shanghai, China). β-actin was used as a housekeeping gene with a 186-bp PCR product for normalization (forward: 5′-TGGCACCCAGCACAATGAA-3′, reverse: 5′-CTAAGTCATAGTCCGCCTAGAAGCA-3′). The following primers were used to amplify ELAVL1 with a 229-bp product (forward: 5′-AGAGCGATCAACACGCTGAA-3′, reverse: 5′-TAAACGCAACCCCTCTGGAC-3′). The RT-qPCR reaction began with a 10-minute denaturation at 95 ℃, followed by 40 cycles of 95 ℃ for 15 s and 60 ℃ for 45 s. The reactions were conducted in triplicates, and the relative quantification was performed using the comparative 2^-ΔΔCT^ method.^22^

### Western Blotting

Whole cell lysates were prepared using the RIPA buffer and 100 mM PMSF at a volume ratio of 100:1. Protein quantification was achieved using the BCA method. Then, 15 μg of protein in each sample was added to the SDS-PAGE using a 12% gel and transferred to PVDF membranes, which were incubated overnight with the corresponding primary antibody. The primary and secondary antibodies used in the western blotting analysis were purchased from HuaBio Co., Ltd. (Hangzhou, China). These included anti-ELAVL1 (Catalog# ET1705-81, 1:2000), anti-CDK4 (Catalog# ET1612-1, 1:1000), anti-CDK6 (Catalog# ET1612-3, 1:2000), and HRP-conjugated β-actin (Catalog# HA601037, 1:5000) antibodies. Subsequently, the HRP-conjugated secondary antibody was incubated, and the chemiluminescent signal was detected using enhanced chemiluminescence reagent.

### Cell Culture and Transfection Using Small Interfering RNAs (siRNAs)

BEAS-2B, A549, NCI-H1299, PC9, H322, and H1650 cells cell lines were sourced from the American Type Culture Collection. Cells were cultured in RPMI 1640 medium supplemented with 10% fetal bovine serum and incubated at 37 °C in a 5% CO_2_-humidified environment. ELAVL1 was knocked down in PC9 using targeted siRNAs commercially synthesized by General Biol (Anhui, China). The sequences of these siRNAs were as follows: si-NC: 5′-UUCUCCGAACGUGUCACGUTT-3′, si-ELAVL1-a: 5′-GAGGUGAUCAAAGACGCCAACUUGU-3′, si-ELAVL1-b: 5′-CCUCGUGGAUCAGACUACAGGU UUG-3′, and si-ELAVL1-c: 5′-CCAACAAGUGCAAAGGGUUUGGCUU-3′. PC9 cells at 60–70% confluence were transfected with siRNAs using Lipofectamine 3000, according to the manufacturer’s protocol (Thermo Fisher Scientific, Waltham, MA, USA). Subsequent experiments were performed 48 h after transfection.

### Cell Proliferation, Migration, Invasion and Apoptosis Assays

The siRNA used in relative cell functional assays were si-ELAVL1-a and si-ELAVL1-c. The Cell Counting Kit-8 (CCK-8) assay was used to assess the proliferative ability of different groups. In short, 48 hours post-transfection, cells from different groups were placed into 96-well plates, with each well containing 5000 cells in 100 μL of complete culture medium. Cells in each well were exposed to 10 μL of CCK-8 solution for 120 min following incubation periods of 6, 24, 48, or 72 hours (Beyotime Biotechnology Co. Ltd., Shanghai, China). Using a microplate spectrophotometer from Invitrogen, the optical density of the mixture was recorded at 450 mm. Cell migration, invasion, and apoptosis were analyzed using wound healing, transwell, and flow cytometry assays, following the methods described in a previous study.^16^

### Statistical Analysis

All data were analyzed using R version 4.2.2, GraphPad Prism 8, and SPSS 24. Student’s two-sided t-test was used to compare the differences between the two groups. Statistical significance was set at *P* < 0.05.

## Results

### Identification of Prognostic DEFGs in LUAD

A total of 1,121 differentially expressed genes (DEGs) were identified between lung adenocarcinoma (LUAD) and normal lung tissue samples (P < 0.05 and |log2 fold change| ≥ 2).Among them, 798 genes were upregulated and 323 were downregulated(Figure 1(a),(b)).Eight ferroptosis-related differentially expressed genes (DEFGs) were obtained from the intersection of 154 ferroptosis-related genes and these 1,121 DEGs(Figure 1(c),(d)).

**Figure 1.**
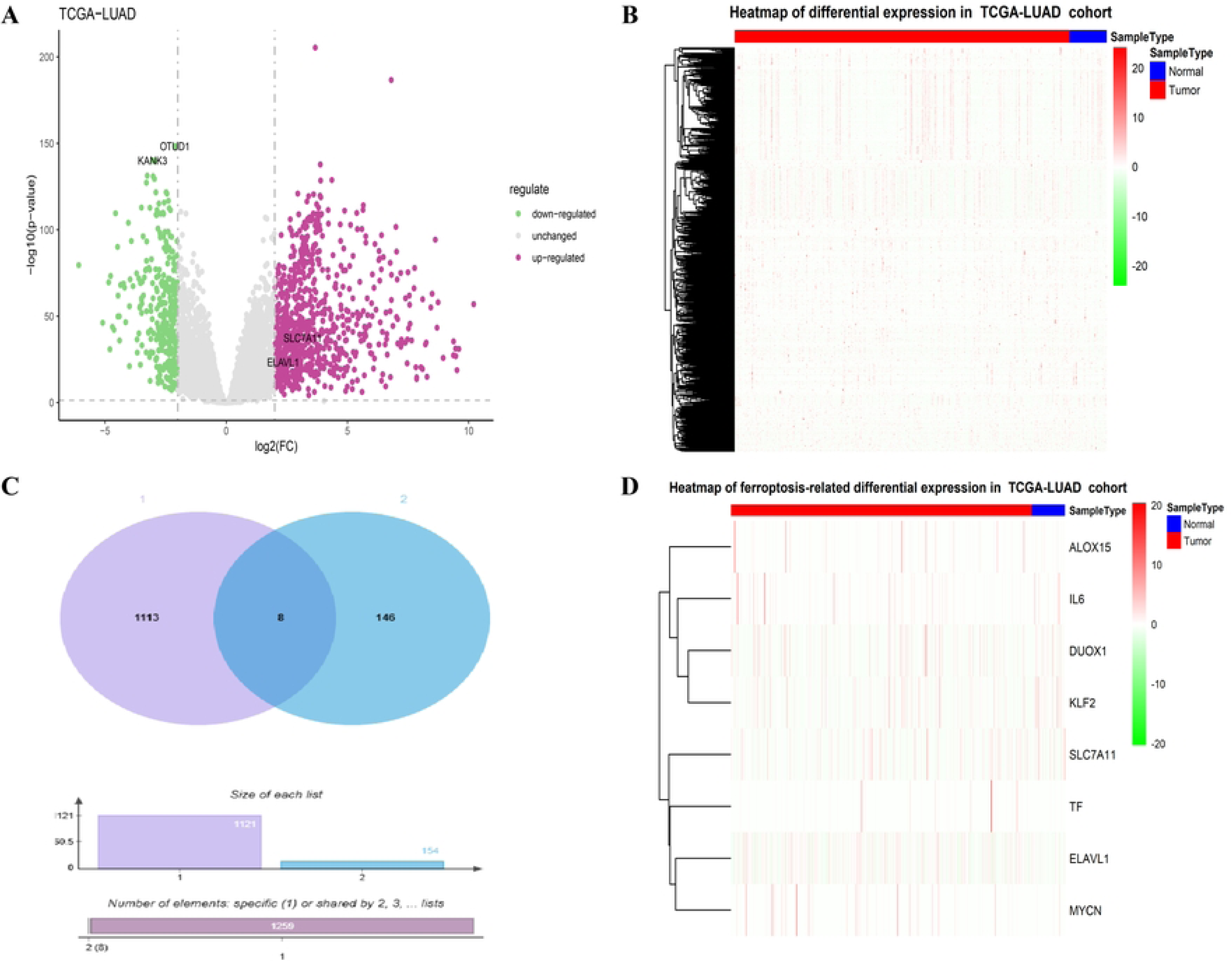
Identification of differentially expressed ferroptosis-related genes. (a, b) Volcano map and heatmap depict the upregulated and downregulated genes between LUAD and adjacent non-tumorous samples in TCGA database. (c) Venn diagram illustrates the intersection of eight genes between the DEG and ferroptosis-related gene sets. (d-e) Heatmap and histogram plot display the expression levels of these eight genes in LUAD and normal lung tissues. \**P* < 0.05 and \*\*\**P* < 0.001 compared with adjacent normal lung tissues.

By analyzing the correlation between the expression of these 8 intersection genes and the clinical data of LUAD patients, we found that patients with high expression of ELAVL1 and SLC7A11 had lower overall survival rates (*P* = 0.0069 for *ELAVL1*, *P* = 0.0094 for *SLC7A11*;Figure 2(a),(b)). However, the expression of six genes, such as KLF2, had no significant relationship with patient survival and prognosis(Figure 2(c)-(h)). Additionally, the mRNA expression levels of ELAVL1 and SLC7A11 were higher in LUAD tissues (*P* = 2.415e-37 for *ELAVL1*, *P* = 3.374e-14 for *SLC7A11*;Figure 1(e)). Thus, high expression of these two genes may indicate a poor prognosis for LUAD patients.

**Figure 2.**
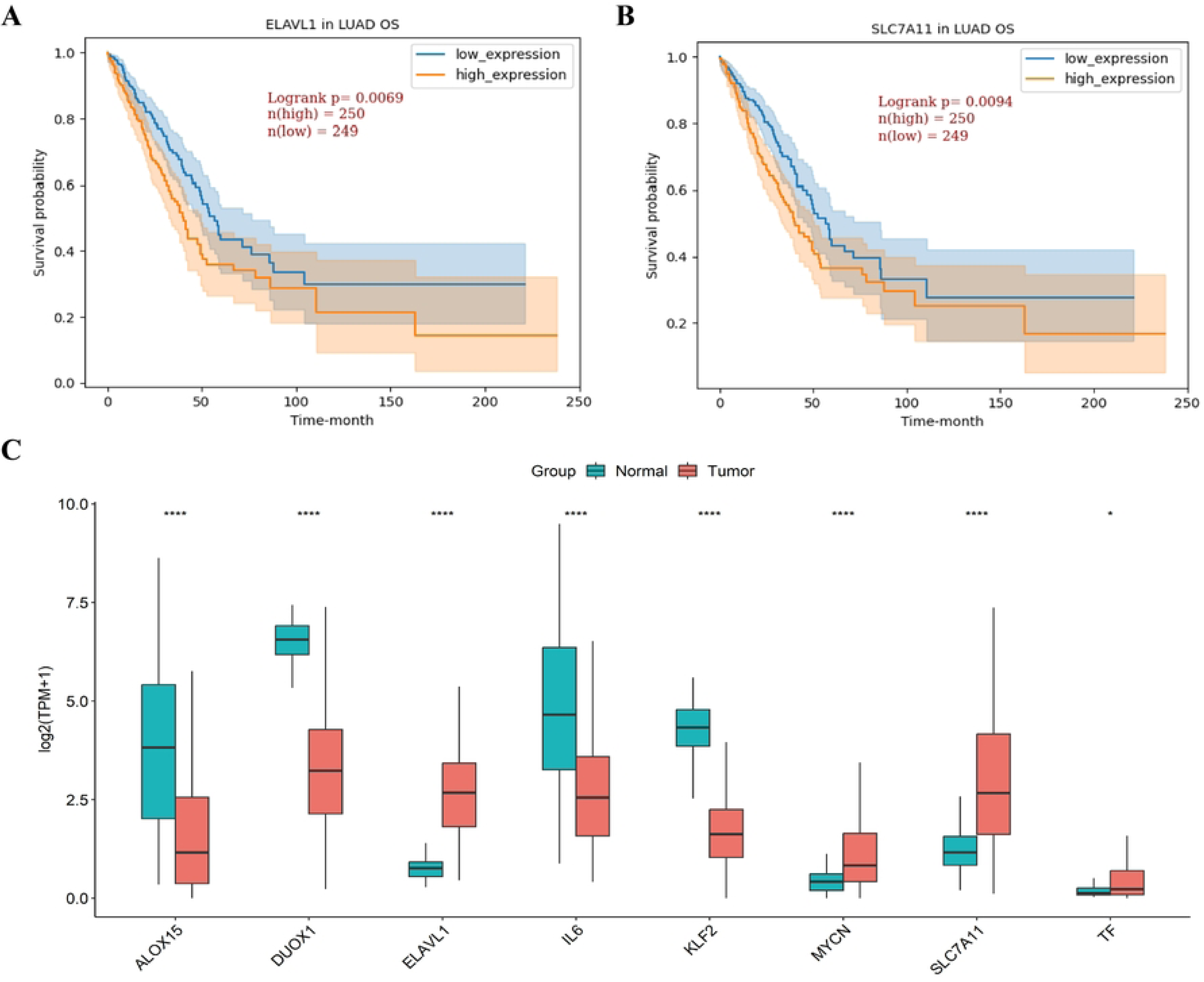
Identification of potential prognostic ferroptosis-related DEGs in LUAD. (a-h) Kaplan–Meier curves with the log-rank test were conducted for the overall survival analysis of hub genes (ELAVL1, SLC7A11, KLF2, DUOX1, TF, IL6, ALOX15, and MYCN).

### Construction and Validation of the Diagnostic Model

To construct and validate the two-gene diagnostic model, 585 samples (526 LUAD samples and 59 normal lung samples) were selected from the TCGA LUAD dataset. These samples were divided into a training set (n = 409, with 362 LUAD samples and 47 normal lung samples) and a testing set (n = 176, with 164 LUAD samples and 12 normal lung samples). The model was constructed on the training set using the scikit-learn package in Python (version 3.8). The binomial classifier achieved an overall accuracy of 0.936 in the cross-validation of the training set (Figure 3(a)), and the optimal parameters were determined as C = 1.234 and gamma = 0.754. On the testing set, the model’s AUC value was 0.9675 (Figure 3(b)), indicating that the two-gene diagnostic model had good specificity for LUAD diagnosis.

**Figure 3.**
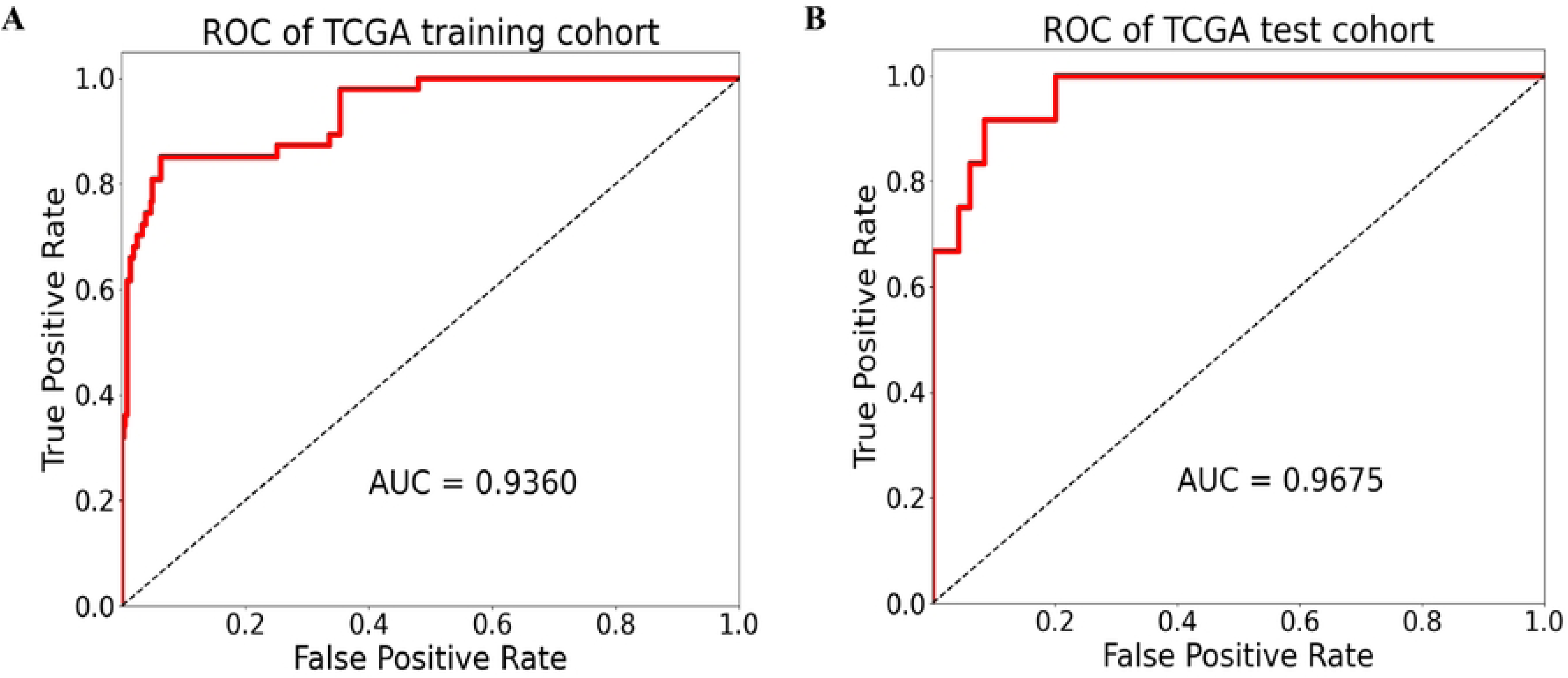
Establishment and validation of the diagnostic model. (a) ROC curve analysis for the efficacy of the SVM diagnostic model based on two hub genes in the training sets. The x-axis indicates the FPR, and the y-axis indicates the TPR. The five-fold cross-validation is represented. The black dotted line denotes the final fitted average. (b) ROC curve analysis for model efficacy based on the testing sets. The x-axis represents the FPR, and the y-axis represents the TPR. SVM, support vector machine; ROC, receiver operating characteristic; FPR, false-positive rate; TPR, true-positive rate.

### Development and Assessment of the Prognostic Model

Data from 508 LUAD patients with OS status were collected from the combined datasets of TCGA and clinical records. A prognostic model was constructed using the formula “Risk score = (0.399×ELAVL1) + (0.105×SLC7A11)” (Table 1). The 508 patients were evenly divided into a training set and a testing set (254 patients each), and patients in each set were classified into high- and low-risk groups based on a median risk score of 2.3433.

According to the distribution of risk scores, outcome status, and profiles of the two-gene signature in each set, the results showed that the mortality rate in the high-risk group was high, and most patients in the low-risk group survived throughout the follow-up period (Figure 4(a)-(c)). The heatmap revealed that the two DEFGs had different expression patterns in different risk groups. The KM survival curves indicated that the overall survival (OS) of high-risk patients was poorer than that of low-risk patients (Figure 4(d)-(f)).

**Figure 4.**
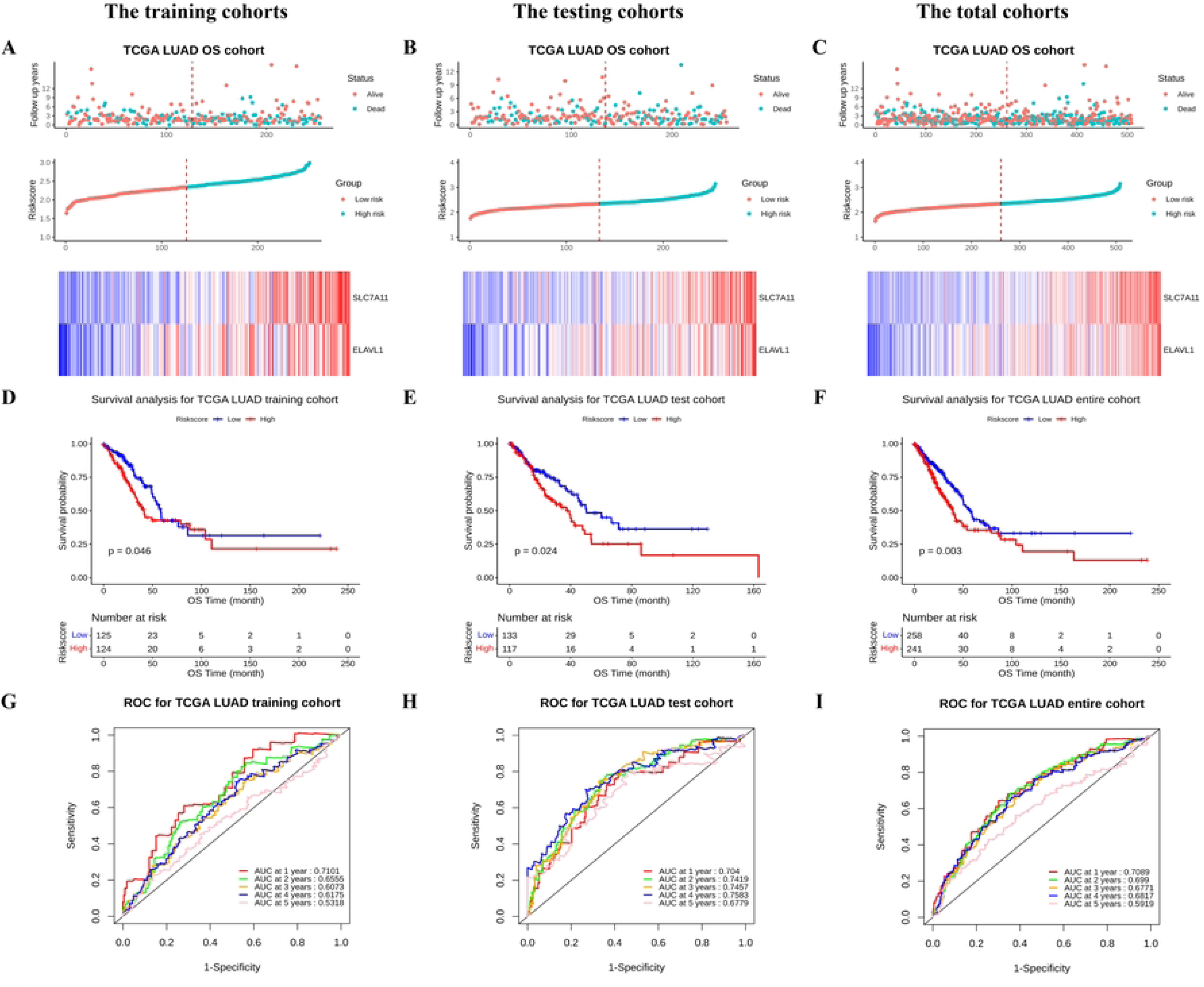
Establishment and validation of the prognostic model for OS of LUAD patients from TCGA database. (a-c) Distribution of risk scores, gene expression levels, and patient relapse status in the training, testing, and total cohorts. (d-f) Kaplan–Meier curves of OS of the low- and high-risk groups. (g-i) ROC curve for the 1-, 2-, 3-, 4-, and 5-year survival prediction by the two-gene signature. The black dotted line represents the median risk score cutoff dividing patients into low- and high-risk groups.

We also evaluated the performance of the two-gene signature in predicting 5-year survival (Figure 4(g)-(i)) and used ROC curve analysis to assess its predictive value. In the training, testing, and total cohorts, the AUC values for 1 - 5-year OS were as follows: for the training cohort, 0.7101, 0.6555, 0.6073, 0.6175, and 0.5318; for the testing cohort, 0.704, 0.7419, 0.7457, 0.7583, and 0.6779; for the total cohort, 0.7089, 0.699, 0.6771, 0.6817, and 0.5919. These results suggest that the prognostic model is effective in predicting the OS risk of LUAD patients.

### Independent Prognostic Value of Two-Gene Signature

Independent prognosis analyses were conducted on clinical characteristics and risk scores in relation to patient survival. Cox regression analyses, both univariate and multivariate, were conducted on the clinical characteristics and risk score of overall samples from TCGA dataset. Univariate cox regression analysis identified significant associations between OS and the following factors: risk score (HR = 1.622; 95% CI [1.203–2.185]; *P* = 0.002), pathological stage (HR = 2.085; 95% CI [1.344–3.234]; *P* = 0.001), tumor stage (HR = 2.209; 95% CI [1.227–3.976]; *P* = 0.008), lymph node metastasis (HR = 2.511; 95% CI [1.861–3.387]; *P* < 0.001), and distant metastasis (HR = 1.997; 95% CI [1.165–3.423]; *P* = 0.0119) as shown in Table 2. Multivariate cox regression analysis indicated significant associations between OS and both the risk score (HR = 1.78; 95% CI [1.247–2.541]; *P* = 0.001) and lymph node metastasis (HR = 12.543; 95% CI [1.793–3.608]; *P* < 0.001). These results validate that this two-ferroptosis gene signature was predictive of survival in the independent validation LUAD cohorts.

### Functional Analysis of the Prognostic Signature

The potential signaling pathways related to the two ferroptosis-associated genes in LUAD were investigated. GO analysis revealed that the DEGs were significantly enriched in biological processes (BPs) associated with cell division, cell cycle, cell growth, and metabolism. Furthermore, the DEGs were enriched in cellular components (CCs) such as chromosome-associated structures, proteasomes, and complexes of fibrillar collagen trimer (Figure 5(a)). The bar plot in Figure 5(b) illustrates the significantly enriched KEGG pathways, including cell cycle, protein digestion and absorption, complement and coagulation cascades, glycosphingolipid biosynthesis (lacto and neolacto series), and ABC transporters. GSEA confirmed the significant enrichments of the cell cycle signaling pathway, T and B cell receptor signaling pathways, glycometabolism-related pathways, proteasome pathway, cancer-associated pathways, and FcγR-mediated phagocytosis signaling pathway in the high-risk group (Figure 6(a)-(h)). In contrast, only ABC transporters were active in the low-risk group (Figure 6(i)).

**Figure 5.**
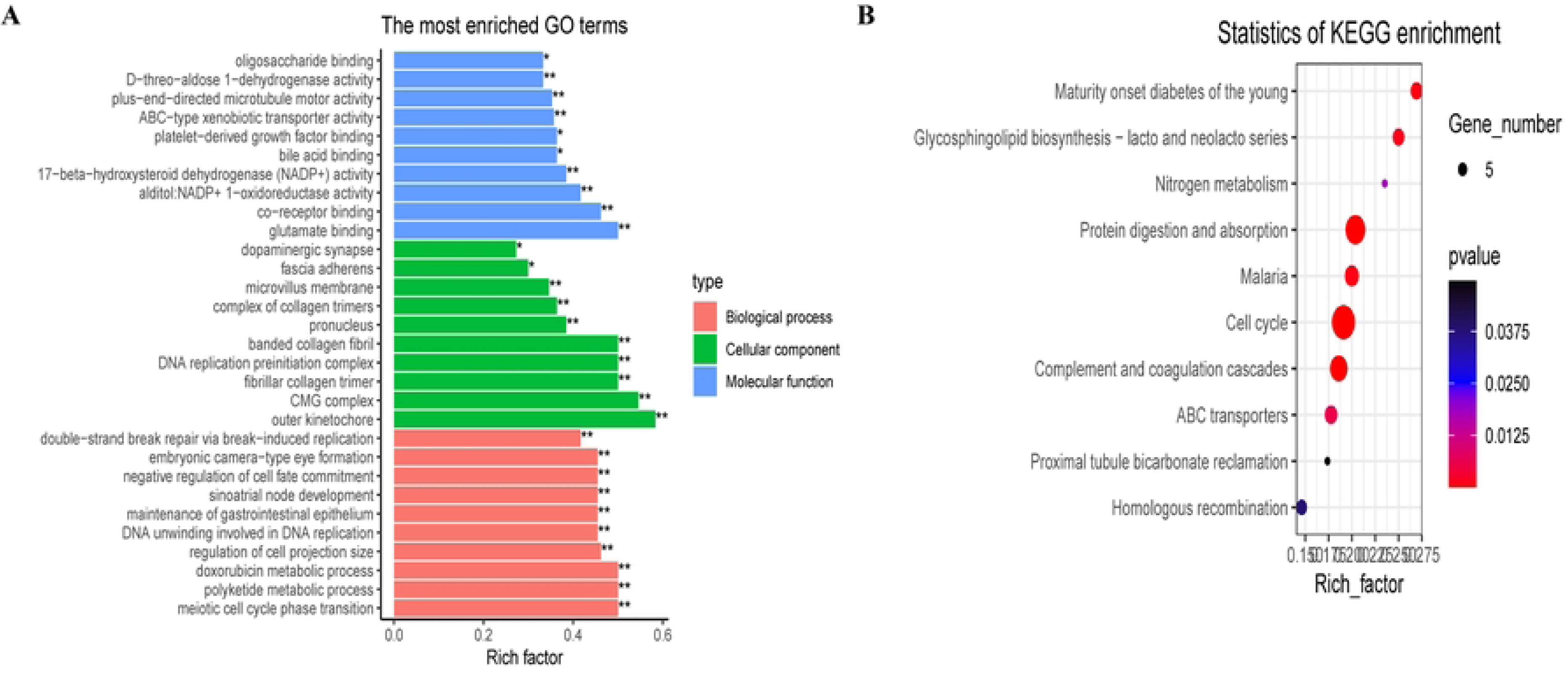
Identification of biological pathways associated with DEGs between LUAD and normal samples from TCGA database. (a) GO analysis of DEGs, including BP, CC, and MF. (b) KEGG analysis of DEGs. GO, Gene Ontology; BP, biological process; CC, cellular component; MF, molecular function; KEGG, Kyoto Encyclopedia of Genes and Genomes.

**Figure 6.**
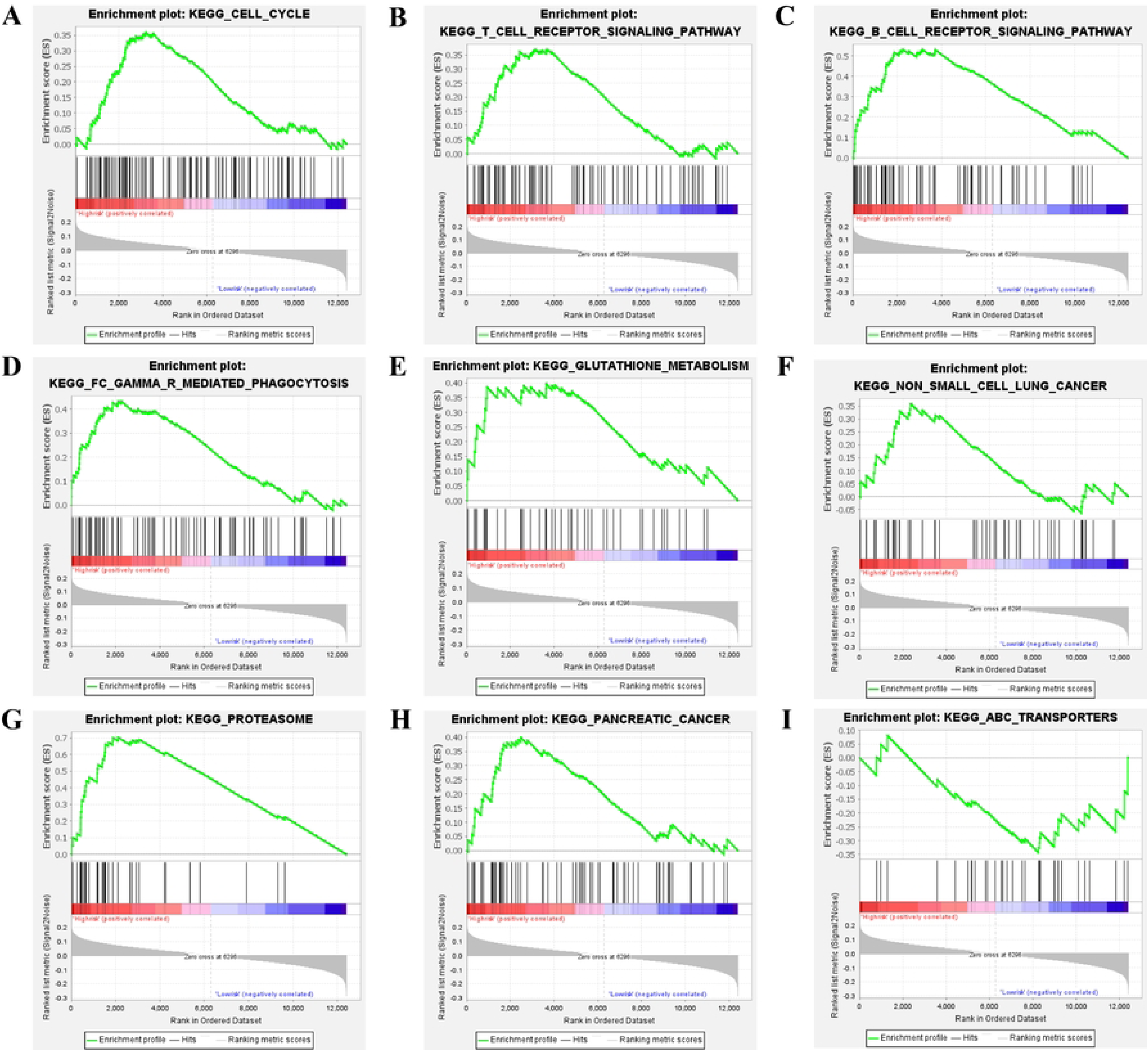
Gene set enrichment analysis of genes correlated with prognostic signature in the high- and low-risk groups. (a-h) Significantly enriched KEGG pathways in the high-risk group. (i) Significantly enriched KEGG pathways in the low-risk group.

### Analysis of Immune Cell Infiltration in the Prognostic Signature

The CIBERSORT algorithm was used to quantify 22 immune cell populations in each LUAD sample, applying a significance threshold of *P* < 0.05 to analyze the association between immune cell infiltration and the prognostic signature. The analysis revealed that resting memory CD4+ T cells were the most abundant infiltrating cells, followed by M2 macrophages, naive B cells, and M0 macrophages (Figure 7(a),(b). Differences in immune cell infiltration were observed between the high- and low-risk groups. Figure 7(c) indicates that the proportion of monocytes was significantly enriched in the low-risk group compared with that in the high-risk group (*P* = 0.0014). A higher proportion of naive B cells was significantly present in the high-risk group (*P* = 0.027).

**Figure 7.**
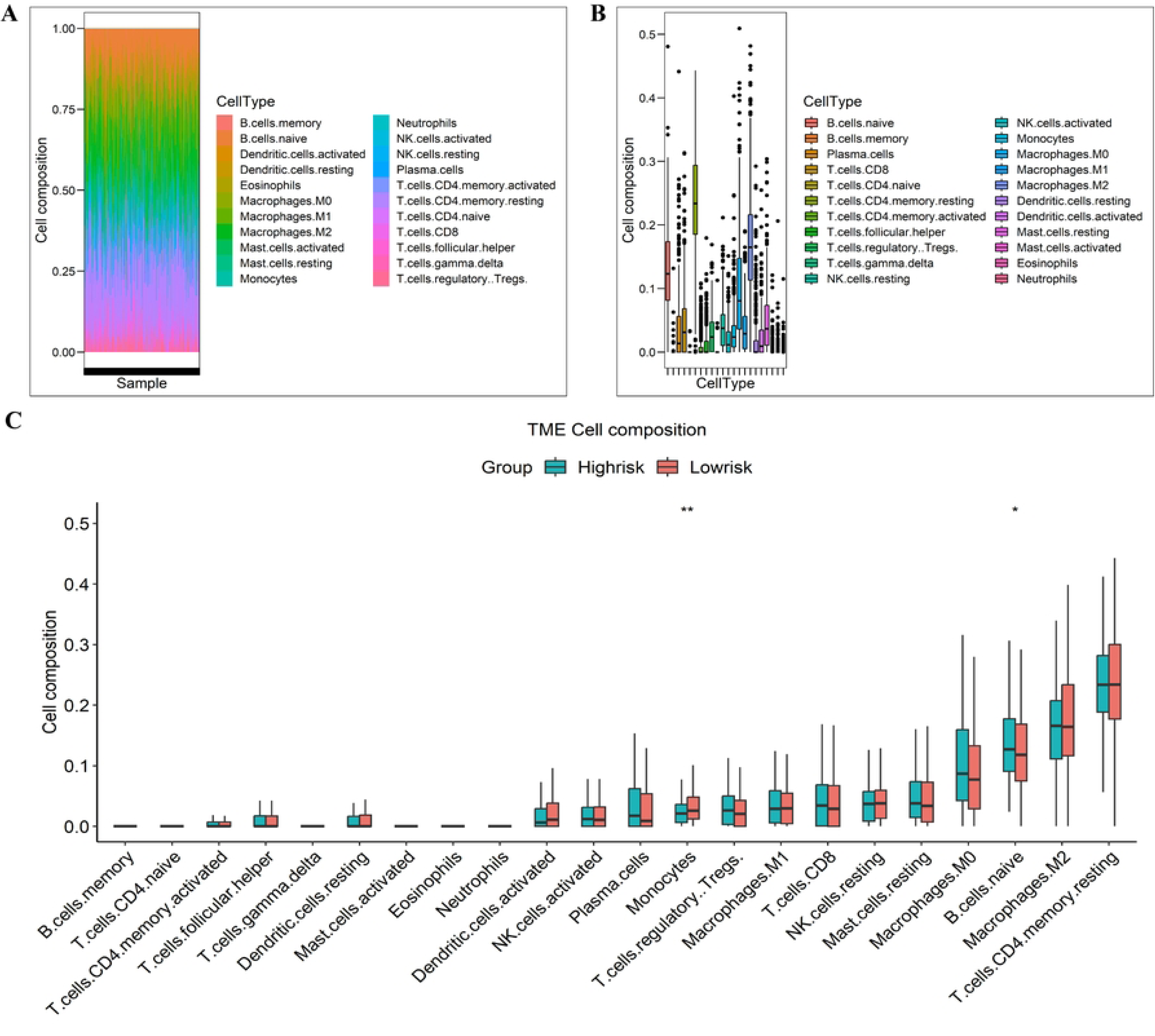
Analysis of immune cell subtypes in LUAD using CIBERSORT. (a) Bar chart displaying the proportion of immune cell subsets. The x-axis depicts the sample names, and the y-axis indicates the percentage of 22 immune cell subsets. (b) Composition of 22 types of immune cells in LUAD tissue samples. (c) Histogram plot of the percentage of immune cells in the high- and risk-score groups. Blue and red represent the high- and low-risk groups, respectively. The x-axis displays the names of 22 immune cell subtypes, and the y-axis represents the corresponding percentage of these immune cell subtypes. \**P* < 0.05 and \*\**P* < 0.01 compared with low-risk group.

### Experimental Verification of Hub Genes through Immunohistochemistry Staining

To further analyze the expression of ELAVL1 and SLC7A11 in patients, we collected normal lung and LUAD tissues (Supplementary Table 1). Immunohistochemical staining revealed significantly higher protein expression levels of ELAVL1 and SLC7A11 in LUAD tissues than in normal lung tissues (Figure 8(a),(b),(c),(e)). In LUAD samples, ELAVL1 was predominantly localized in the nucleus, displaying strong positive staining, whereas SLC7A11 was prominently observed in the cytoplasm with moderate to strong positive staining. Furthermore, we investigated the relationship between the expression profiles of these two hub prognostic genes and the pathological stage. Our analysis demonstrated that only the protein expression level of ELAVL1 increased with the progression of LUAD (Figure 8(d)). Conversely, the protein expression of SLC7A11 did not show statistically significant differences between stages II and III (Figure 8(f)). Research on genes regulating tumor progression in LUAD, except for *SLC7A11*, is currently limited. Taken together, these data suggest that *ELAVL1* may be a potential gene associated with ferroptosis prognosis in the progression of LUAD.

**Figure 8.**
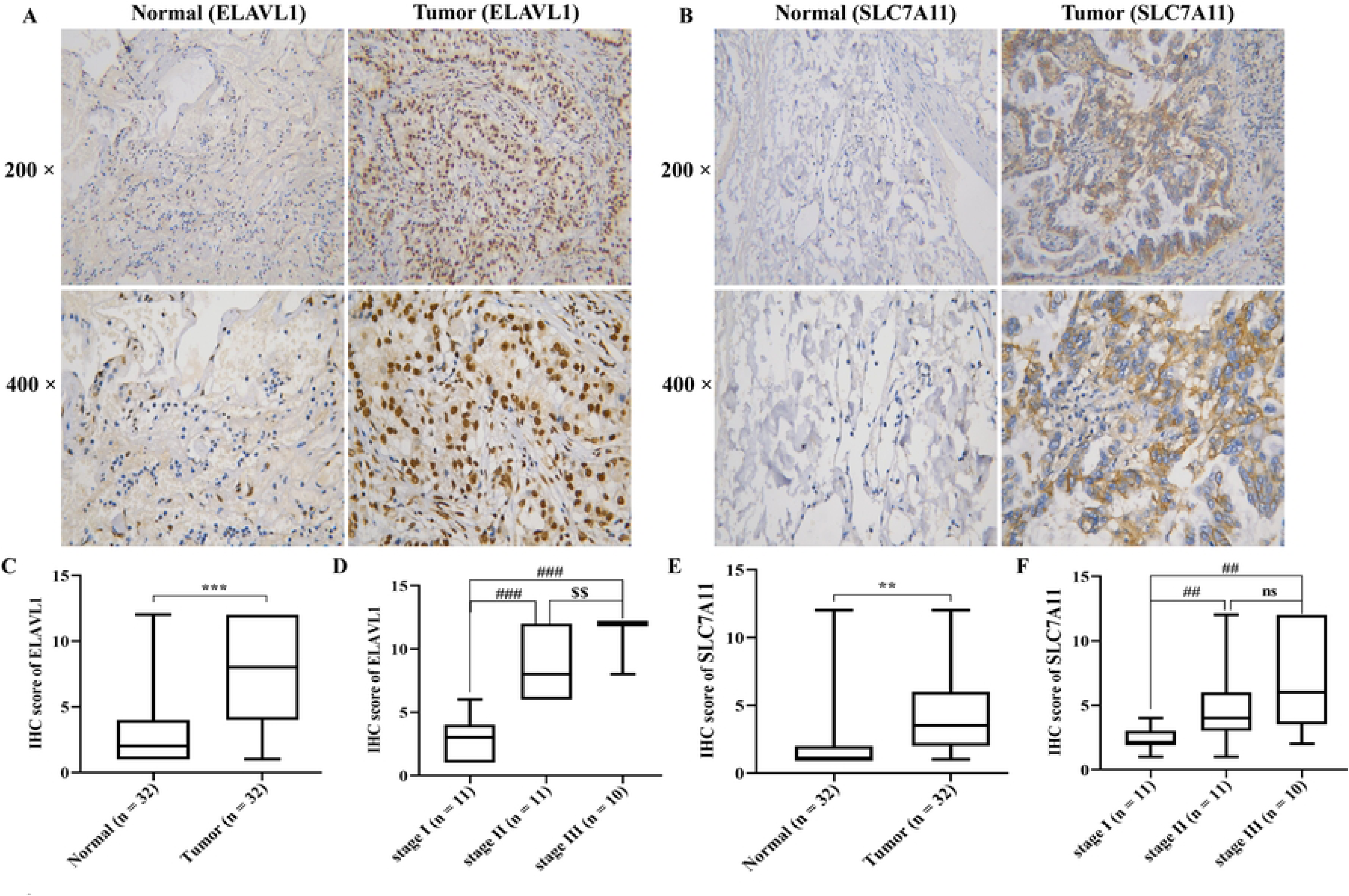
Expression levels of SLC7A11 and ELAVL1 in adjacent normal lung and LUAD tissues. (a, b) Representative immunohistochemistry staining images of ELAVL1 and SLC7A11 in adjacent normal lung and LUAD tissues. (c, e) Statistical results of ELAVL1 and SLC7A1 expression in 30 pairs of LUAD and adjacent normal lung tissues. \*\**P* < 0.01 and \*\*\**P* < 0.001 compared with adjacent normal lung tissues. (d) Correlation between ELAVL1 expression level and various pathological stages. ^###^*P* < 0.001 compared with stage I; ^$$^*P* < 0.01 compared with stage II. (f) Correlation between SLC7A11 expression level and various pathological stages. ns, not significant; ^##^*P* < 0.001 compared with stage I.

### Silencing ELAVL1 Promotes the Malignant Phenotype of LUAD in Vitro

To further evaluate the specific role of ELAVL1 in LUAD, we examined its expression levels. The expression of ELAVL1 in the PC9 cells was higher than that in the other lung cancer cell lines and BEAS-2B cells (Figure 9(a),(b)). Therefore, the PC9 cell line was selected for subsequent functional assays. The efficiency of ELAVL1 knockdown was verified through RT-qPCR and western blotting (Figure 9(c),(d)). Western blotting analysis showed the inhibition of cell cycle-associated proteins (Figure 9(e)), suggesting that ELAVL1 predominantly regulates LUAD progression through cell cycle modulation. Subsequently, we investigated the potential functions of ELAVL1 in PC9 cells through cell proliferation, migration, and invasion assays. The CCK-8 assay revealed a reduced proliferative rate in PC9 cells upon ELAVL1 knockdown (Figure 9(f)).Moreover, wound healing and transwell assays demonstrated significantly impaired cell migration and invasion rates upon ELAVL1 knockdown (Figure 9(g)-(i)). Flow cytometry analysis further indicated that the apoptosis induced by ELAVL1 knockdown might contribute to the inhibition of proliferation (Figure 9(j),(k)).Based on these findings, we hypothesize that the upregulated oncogene *ELAVL1* is involved in LUAD proliferation, migration, and invasion by influencing ferroptosis levels and cell cycle regulation. However, further investigations are warranted to comprehensively elucidate the underlying molecular mechanisms involved in these processes.

**Figure 9.**
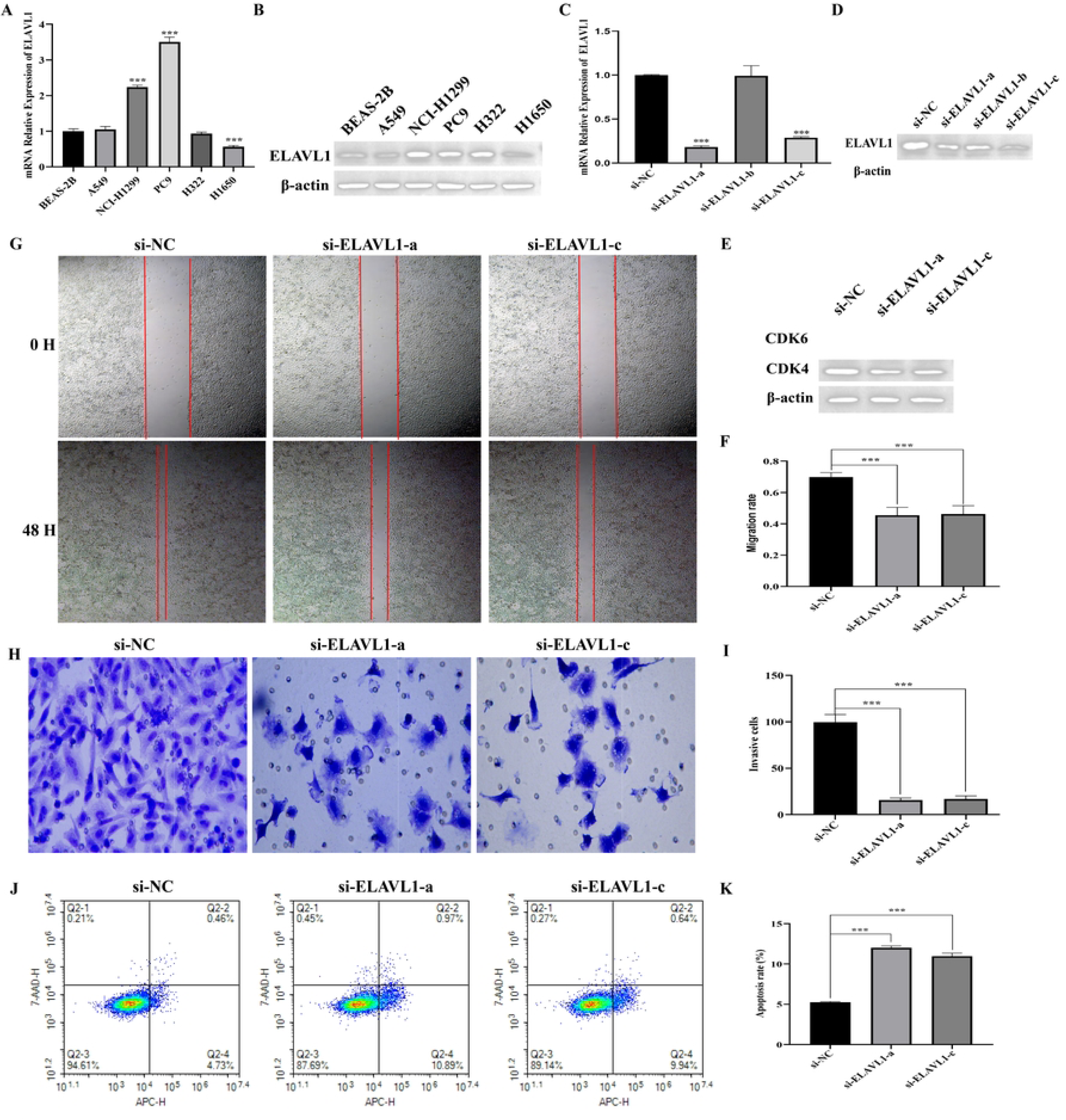
Knockdown of ELAVL1 inhibits the tumor progression of LUAD in vitro. (a, b) Expression levels of ELAVL1 in immortalized human bronchial epithelial mesothelial and NSCLC cell lines determined through RT-qPCR and western blotting. β-actin served as the internal reference. Data represent the mean ± SD of three independent experiments. \*\*\**P* < 0.001 compared with BEAS-2B. (c, d) Knockdown efficiency of ELAVL1 si-RNAs (si-NC, si-ELAVL1-a, si-ELAVL1-b, and si-ELAVL1-c) in PC9 cell lines confirmed through RT-qPCR and western blotting. β-actin served as the internal reference. Data represent the mean ± SD of three independent experiments. \*\*\**P* < 0.001 compared with si-NC. (e) Protein expression levels of several cell cycle regulators in PC9 after knocking down ELAVL1 determined through western blotting. β-actin served as the internal reference. Data represent the mean ± SD of five independent experiments. (f) Effect of ELAVL1 knockdown on PC9 proliferation determined through CCK-8 assay. Data represent the mean ± SD of five independent experiments. \*\*\**P* < 0.001 compared with si-NC. (g) Representative images of the wound healing assay and statistical analysis of the wound healing assay results after knockdown of ELAVL1 in PC9. (h, i) Representative images of the transwell assay and statistical analysis after knockdown of ELAVL1 in PC9. Data represent the mean ± SD of three independent experiments. \*\*\**P* < 0.001 compared with si-NC. (j, k) Flow cytometry assay (representative images presented) and statistical analysis used to confirm apoptosis induced by ELAVL1 knockdown. Data represent the mean ± SD of three independent experiments. \*\*\**P* < 0.001 compared with si-NC.

## Discussion

With the increasing popularity of high-resolution CT scanning and improved patient health awareness, the detection rate of pulmonary nodules has increased.^23^ Nonetheless, accurately differentiating between benign and malignant lung nodules associated with LUAD remains a significant challenge for clinicians. Currently, most LUAD patients are diagnosed at an advanced stage, and the clinical outcomes of LUAD treatment remain unsatisfactory. Various factors, including clinical factors and genes, influence the prognosis of LUAD patients.^24, 25^ Consequently, investigating diagnostic and prognostic markers for LUAD is crucial. Mounting data suggests that ferroptosis, a recently identified form of programmed cell death, plays a crucial role in carcinogenesis and the efficacy of cancer therapies.^6, 26^ However, existing ferroptosis-related studies in LUAD have predominantly focused on the role of potential ferroptosis biomarkers in inducing ferroptosis with well-established inducers. Consequently, comprehensive and systematic analysis of ferroptosis in accurate diagnosis, individualized treatment, and prognosis prediction for LUAD are currently lacking. In this study, we identified two hub genes, SLC7A11 and ELAVL, by intersecting DEGs in LUAD with genes associated with OS in LUAD.

Diagnosis and prognostic prediction represent crucial steps in the management of LUAD patients. To identify potential biomarkers for the diagnosis and prognosis of LUAD, we constructed a diagnostic model using *SLC7A11* and *ELAVL*. The two-gene signature exhibited excellent diagnostic ability, as evidenced by the AUC values when applied to LUAD and normal tissue samples in TCGA database. Subsequently, we developed a prognostic model to provide a robust indicator for evaluating OS in LUAD patients, thereby illustrating the impact of the prognostic signature. Furthermore, the high expression levels of SLC7A11 and ELAVL observed in LUAD tissues were consistent with the data from TCGA database. These findings suggest a potential association between the expression alterations of SLC7A11 and ELAVL and the prognosis of LUAD patients.

*SLC7A11* encodes a chloride-dependent sodium-independent cystine-glutamate antiporter known as “system Xc-” or “xCT.”^27^ It plays a role in regulating synaptic activity by stimulating extra-synaptic receptors and facilitating nonvesicular glutamate release.^28^ SLC7A11 is overexpressed in several cancer types, including glioma, liver carcinoma, NSCLC, and triple-negative breast cancer, and it serves as an independent prognostic factor.^29–33^ Studies have reported that high expression of SLC7A11 predicts an advanced clinical pathological stage in LUAD patients.^34^ In clinical practice, the pathological stage is considered a key factor for predicting the clinical outcome of LUAD patients, and its extent is strongly associated with distant metastasis.^35, 36^ Our results further confirmed the association between increased expression levels of SLC7A11 and the progression of LUAD in pathology. ELAVL1 is an RNA-binding protein that binds to AU- and U-rich element regions of target messenger RNAs, thereby regulating their stability and promoting gene expression.^37–39^ This protein, known to be involved in various cellular processes such as angiogenesis, apoptosis, and inflammation, plays a crucial role.^38–40^ Previous studies have demonstrated elevated levels of ELAVL1 in the diabetic heart, suggesting its involvement in cardiac pyroptosis and ferroptosis in liver fibrosis.^41–43^ Additionally, several studies have reported a tumor-promoting effect of ELAVL1 in different cancer types. Barbisan et al. and Melling et al. reported an upregulation of ELAVL1 in prostate cancer, indicating that ELAVL1 promotes the development of prostate cancer.^44, 45^ Kanzaki et al. demonstrated the high expression of ELAVL1 in HCC tissues and the reduced proliferative activity of HepAD38 cells upon ELAVL1 knockdown.^46^ Similarly, in our study suppressed proliferative, motile, invasive, and migratory abilities of LUAD cells were observed upon ELAVL1 knockdown in PC9 cells. Moreover, we found a significant increase in the expression of ELAVL1 protein with the increase in pathological grade from 1 to 3 in LUAD tissues. Based on the aforementioned evidence, we propose that SLC7A11 and ELAVL could be important targets for tumor treatment. Additionally, our study is the first to report that ELAVL1 is essential for the progression of LUAD, both in vitro and in clinical tissues. Further experimental studies are necessary to uncover the detailed role of ELAVL1 in LUAD.

For clinical application, effective biomarkers should accurately predict prognosis for patients, distinguish between patients with different risks, and thus assist clinicians in formulating the most reasonable treatment plans promptly.^47^ In our study, the prognostic signature successfully stratified LUAD patients into high- and low-risk groups, revealing statistically significant differences in OS outcomes. Subsequently, we systematically analyzed the patient risk score and clinical information in TCGA (including pathological and TNM stages) for OS using univariate and multivariate Cox regression analysis. The results indicated that our signature could serve as an independent prognostic factor for predicting the prognosis of patients.

During the process of constructing and validating the prognostic model, we observed distinct immune statuses among LUAD patients, as evidenced by expression patterns of the DEFGs. Patients with a more active immune status exhibited better prognosis. Previous studies have highlighted the significance of TIICs as crucial components of the tumor microenvironment, capable of effectively predicting patient prognosis.^48^ Our findings revealed that LUAD samples primarily comprised resting memory CD4+ T cells, M2 macrophages, naive B cells, and M0 macrophages in terms of immune cell composition, suggesting their involvement in LUAD development and progression. Numerous studies have demonstrated that elevated levels of B cells, T cells, and macrophages, including naive B cells, resting memory CD4+ T cells, M0 macrophages, and M2 macrophages, are associated with worse OS in various cancer types.^49–52^ Additionally, we observed differences in immune cell infiltration between the high- and low-risk groups, indicating significant variations in immune cell composition between these patient cohorts. Specifically, high-risk patients exhibited a positive correlation with immune infiltration of naive B cells. Naive B cells, characterized by their lack of exposure to antigens,^53–55^ contribute to the inactivation of naive T cells, affecting T cell function and promoting immune evasion by tumor cells. This suggests that a high level of naive B cells is closely associated with poor survival in patients with a high-risk score. Furthermore, our results indicated a low level of monocytes in the high-risk group. Previous studies have highlighted the crucial role of monocytes in the immune system, where their activation represents a critical step in mounting immune responses.^56^ In the early stages of tumor progression, the recruitment of monocytes is observed in various cancer types, and they contribute to the direct elimination of malignant cells through cytokine-mediated cell death and phagocytosis.^57, 58^ Therefore, a low level of monocytes serves as a poor prognostic factor in LUAD patients with a high-risk score. Overall, the ferroptosis-related gene signature, incorporating *SLC7A11* and *ELAVL1*, not only holds potential as a prognostic predictor in lung adenocarcinoma but may also function as a significant regulator of tumor immunity. Nevertheless, additional validation is necessary to comprehensively understand the role of our signature in predicting the immunotherapeutic response in LUAD patients.

Although extensive research has focused on investigating the mechanism and clinical application of ferroptosis, particularly in the field of tumor therapy, the underlying mechanisms governing tumor susceptibility to ferroptosis still require further clarification. Initially, KEGG and GO enrichment analyses of the DEGs in LUAD revealed their involvement in various signaling pathways, including the cell cycle, DNA replication, and the p53 pathway. Furthermore, GSEA was performed to examine the pathways associated with the two-gene signature. Categorizing the samples based on the risk score into high- and low-risk groups demonstrated that the high-risk group exhibited enrichment in immune-related signaling pathways and cell cycle signaling compared with the low-risk group. Previous studies have highlighted a strong interaction between ferroptosis and immunity, playing a critical role in tumor initiation, progression, and treatment. Wang et al. first discovered that immunotherapy-activated CD8+ T cells induced ferroptotic behaviors in ovarian tumor cells, indicating a strong link between ferroptosis and anti-tumor immunity.^58, 59^ However, the specific underlying mechanisms that induce ferroptotic behaviors in tumor cells and enhance anti-tumor immune responses remain unclear. In our study, we observed indications that ferroptosis influences the immune status of LUAD through the TCR and BCR signaling pathways. Additionally, dysregulation of cell cycle-related pathways may result from imbalanced ferroptosis. Abnormal expression of cell cycle-regulating molecules may contribute to distinct ferroptosis-related risks in LUAD patients.^4^ The key cell cycle regulators, CDK4 and CDK6, were downregulated after ELAVL1 knockdown in PC9 cells. Based on these findings, we postulate that the upregulation of the two genes may induce alterations in TCR, BCR, and cell cycle regulator contents during the development of LUAD. Nonetheless, the intricate relationships and regulatory mechanisms among ferroptosis, cell cycle, and immune responses necessitate further elucidation.

## Conclusions

We constructed a diagnostic model based on two DEFGs (*SLC7A11* and *ELAVL1*) that demonstrated relatively high diagnostic efficiency for LUAD. Additionally, a novel prognostic model was developed, independently associated with OS, indicating its potential application in predicting the OS of LUAD patients. Moreover, this study revealed the crucial role of the interaction among ferroptosis, cell cycle, and immunity in the development of LUAD. It provides new insights into the investigation of molecular mechanisms and targeted therapies, such as ELAVL1, for LUAD.

## Data Availability

Data used in this study include third-party data from publicly accessible repositories (TCGA: https://portal.gdc.cancer.gov/ GEO: https://www.ncbi.nlm.nih.gov/geo/)

http://www.zhounan.org/ferrdb/

https://www.ncbi.nlm.nih.gov/geo/

https://portal.gdc.cancer.gov/

## Acknowledgments

This study was supported by the Medical Scientific Research Foundation of Guangdong Province of China (No. B2021435), the Guangdong Province Administration of Traditional Chinese Medicine Research Project (No. 20222288).The Research Talent Start-up Project of The First People’s Hospital of Zhaoqing City Funded Foundation (No: YJJ-2020-02-06).

## Supplementary materials

Table S1: Clinical data of the thirty-two patients.

## Notes

### Competing Interest Statement

The authors have declared no competing interest.

### Funding Statement

Yes

### Author Declarations

The research was approved by the Ethics Committee of The First People's Hospital of Zhaoqing City (Approval No.: Ethical review of The First People's Hospital of Zhaoqing City 2021-06-06).

## References

1. Jemal A, Siegel R, Xu J, et al. Cancer statistics, 2010. CA Cancer J Clin. 2010;60(5):277–300.

2. Shukla S, Evans JR, Malik R, et al. Development of a RNA-Seq Based Prognostic Signature in Lung Adenocarcinoma. J Natl Cancer Inst. 2017;109(1):200.

3. Siegel RL, Miller KD, Wagle NS, et al. Cancer statistics, 2023. CA Cancer J Clin. 2023;73(1):17–48.

4. Tian Q, Zhou Y, Zhu L, et al. Development and Validation of a Ferroptosis-Related Gene Signature for Overall Survival Prediction in Lung Adenocarcinoma. Front Cell Dev Biol. 2021;9:684259.

5. Yagoda N, von Rechenberg M, Zaganjor E, et al. RAS-RAF-MEK-dependent oxidative cell death involving voltage-dependent anion channels. Nature. 2007;447(7146):864–868.

6. Dixon SJ, Lemberg KM, Lamprecht MR, et al. Ferroptosis: an iron-dependent form of nonapoptotic cell death. Cell. 2012;149(5):1060–1072.

7. Jiang M, Qiao M, Zhao C, et al. Targeting ferroptosis for cancer therapy: exploring novel strategies from its mechanisms and role in cancers. Transl Lung Cancer Res. 2020;9(4):1569–1584.

8. Ren JX, Sun X, Yan XL, et al. Ferroptosis in Neurological Diseases. Front Cell Neurosci. 2020;14:218.

9. Friedmann Angeli JP, Schneider M, Proneth B, et al. Inactivation of the ferroptosis regulator Gpx4 triggers acute renal failure in mice. Nat Cell Biol. 2014;16(12):1180–1191.

10. Li J, Cao F, Yin HL, et al. Ferroptosis: past, present and future. Cell Death Dis. 2020;11(2):88.

11. Wang WJ, Ling YY, Zhong YM, et al. Ferroptosis-Enhanced Cancer Immunity by a Ferrocene-Appended Iridium(III) Diphosphine Complex. Angew Chem Int Ed Engl. 2022;61(16):202115247.

12. Jia CL, Yang F, Li R. Prognostic Model Construction and Immune Microenvironment Analysis of Breast Cancer Based on Ferroptosis-Related lncRNAs. Int J Gen Med. 2021;14:9817–9831.

13. Lu Y, Qin H, Jiang B, et al. KLF2 inhibits cancer cell migration and invasion by regulating ferroptosis through GPX4 in clear cell renal cell carcinoma. Cancer Lett. 2021;522:1–13.

14. Yang F, Xiao Y, Ding JH, et al. Ferroptosis heterogeneity in triple-negative breast cancer reveals an innovative immunotherapy combination strategy. Cell Metab. 2023;35(1):84–100.

15. Zhang Y, Guo R, Li J, et al. Research progress on the occurrence and therapeutic mechanism of ferroptosis in NSCLC. Naunyn Schmiedebergs Arch Pharmacol. 2022;395(1):1–12.

16. Chen X, Hu S, Han Y, et al. Ferroptosis-related STEAP3 acts as predictor and regulator in diffuse large B cell lymphoma through immune infiltration. Clin Exp Med. 2023;23(6):2601–2617

17. Goe SA. Predicting Breast Cancer Classification Using Various Machine Learning Classification Algorithm. International Journal for Modern Trends in Science and Technology. 2020;6(12):282–285.

18. Halloran JT, Rocke DM. A Matter of Time: Faster Percolator Analysis via Efficient SVM Learning for Large-Scale Proteomics. J Proteome Res. 2018;17(5):1978–1982.

19. Zhu K, Xiaoqiang L, Deng W, et al. Development and validation of a novel lipid metabolism-related gene prognostic signature and candidate drugs for patients with bladder cancer. Lipids Health Dis. 2021;20(1):146.

20. Newman AM, Liu CL, Green MR, et al. Robust enumeration of cell subsets from tissue expression profiles. Nat Methods. 2015;12(5):453–457.

21. Livak KJ, Schmittgen TD. Analysis of relative gene expression data using real-time quantitative PCR and the 2(-Delta Delta C(T)) Method. Methods. 2001;25(4):402–408.

22. Guo T, Zhao S, Li Z, et al. Elevated MACC1 expression predicts poor prognosis in small invasive lung adenocarcinoma. Cancer Biomark. 2018;22(2):301–310.

23. Cai Q, Zhang P, He B, et al. Identification of diagnostic DNA methylation biomarkers specific for early-stage lung adenocarcinoma. Cancer Genet. 2020;246-247:1–11.

24. Ricciuti B, Arbour KC, Lin JJ, et al. Diminished Efficacy of Programmed Death-(Ligand)1 Inhibition in STK11- and KEAP1-Mutant Lung Adenocarcinoma Is Affected by KRAS Mutation Status. J Thorac Oncol. 2022;17(3):399–410.

25. Wohlhieter CA, Richards AL, Uddin F, et al. Concurrent Mutations in STK11 and KEAP1 Promote Ferroptosis Protection and SCD1 Dependence in Lung Cancer. Cell Rep. 2020;33(9):108444.

26. Hassannia B, Vandenabeele P, Vanden Berghe T. Targeting Ferroptosis to Iron Out Cancer. Cancer Cell. 2019;35(6):830–849.

27. Lofthouse EM, Manousopoulou A, Cleal JK, et al. N-acetylcysteine, xCT and suppression of Maxi-chloride channel activity in human placenta. Placenta. 2021;110:46–55.

28. Koppula P, Zhang Y, Zhuang L, et al. Amino acid transporter SLC7A11/xCT at the crossroads of regulating redox homeostasis and nutrient dependency of cancer. Cancer Commun (Lond). 2018;38(1):12.

29. Savaskan NE, Heckel A, Hahnen E, et al. Small interfering RNA-mediated xCT silencing in gliomas inhibits neurodegeneration and alleviates brain edema. Nat Med. 2008;14(6):629–632.

30. Zhang L, Huang Y, Ling J, et al. Overexpression of SLC7A11: a novel oncogene and an indicator of unfavorable prognosis for liver carcinoma. Future Oncol. 2018;14(10):927–936.

31. Timmerman LA, Holton T, Yuneva M, et al. Glutamine sensitivity analysis identifies the xCT antiporter as a common triple-negative breast tumor therapeutic target. Cancer Cell. 2013;24(4):450–465.

32. Ji X, Qian J, Rahman SMJ, et al. xCT (SLC7A11)-mediated metabolic reprogramming promotes non-small cell lung cancer progression. Oncogene. 2018;37(36):5007–5019.

33. Robert SM, Buckingham SC, Campbell SL, et al. SLC7A11 expression is associated with seizures and predicts poor survival in patients with malignant glioma. Sci Transl Med. 2015;7(289):86.

34. Qian L, Wang F, Lu SM, et al. A Comprehensive Prognostic and Immune Analysis of Ferroptosis-Related Genes Identifies SLC7A11 as a Novel Prognostic Biomarker in Lung Adenocarcinoma. J Immunol Res. 2022;2022:1951620.

35. Chansky K, Detterbeck FC, Nicholson AG, et al. The IASLC Lung Cancer Staging Project: External Validation of the Revision of the TNM Stage Groupings in the Eighth Edition of the TNM Classification of Lung Cancer. J Thorac Oncol. 2017;12(7):1109–1121.

36. Feng SH, Yang ST. The new 8th TNM staging system of lung cancer and its potential imaging interpretation pitfalls and limitations with CT image demonstrations. Diagn Interv Radiol. 2019;25(4):270–279.

37. Simone LE, Keene JD. Mechanisms coordinating ELAV/Hu mRNA regulons. Curr Opin Genet Dev. 2013;23(1):35–43.

38. Wang J, Guo Y, Chu H, et al. Multiple functions of the RNA-binding protein HuR in cancer progression, treatment responses and prognosis. Int J Mol Sci. 2013;14(5):10015–10041.

39. Levy NS, Chung S, Furneaux H, et al. Hypoxic stabilization of vascular endothelial growth factor mRNA by the RNA-binding protein HuR. J Biol Chem. 1998;273(11):6417–6423.

40. Srikantan S, Gorospe M. HuR function in disease. Front Biosci (Landmark Ed). 2012;17(1):189–205.

41. Dong R, Chen P, Polireddy K, et al. An RNA-Binding Protein, Hu-antigen R, in Pancreatic Cancer Epithelial to Mesenchymal Transition, Metastasis, and Cancer Stem Cells. Mol Cancer Ther. 2020;19(11):2267–2277.

42. Mao G, Mu Z, Wu DA. Exosomal lncRNA FOXD3-AS1 upregulates ELAVL1 expression and activates PI3K/Akt pathway to enhance lung cancer cell proliferation, invasion, and 5-fluorouracil resistance. Acta Biochim Biophys Sin (Shanghai). 2021;53(11):1484–1494.

43. Shi J, Guo C, Ma J. CCAT2 enhances autophagy-related invasion and metastasis via regulating miR-4496 and ELAVL1 in hepatocellular carcinoma. J Cell Mol Med. 2021;25(18):8985–8996.

44. Barbisan F, Mazzucchelli R, Santinelli A, et al. Overexpression of ELAV-like protein HuR is associated with increased COX-2 expression in atrophy, high-grade prostatic intraepithelial neoplasia, and incidental prostate cancer in cystoprostatectomies. Eur Urol. 2009;56(1):105–112.

45. Melling N, Taskin B, Hube-Magg C, et al. Cytoplasmic accumulation of ELAVL1 is an independent predictor of biochemical recurrence associated with genomic instability in prostate cancer. Prostate. 2016;76(3):259–272.

46. Kanzaki H, Chiba T, Kaneko T, et al. The RNA-Binding Protein ELAVL1 Regulates Hepatitis B Virus Replication and Growth of Hepatocellular Carcinoma Cells. Int J Mol Sci. 2022;23(14).

47. Song C, Guo Z, Yu D, et al. A Prognostic Nomogram Combining Immune-Related Gene Signature and Clinical Factors Predicts Survival in Patients With Lung Adenocarcinoma. Front Oncol. 2020;10:1300.

48. Angelo SP, Shoushtari AN, Keohan ML, et al. Combined KIT and CTLA-4 Blockade in Patients with Refractory GIST and Other Advanced Sarcomas: A Phase Ib Study of Dasatinib plus Ipilimumab. Clin Cancer Res. 2017;23(12):2972–2980.

49. Iglesia MD, Parker JS, Hoadley KA, et al. Genomic Analysis of Immune Cell Infiltrates Across 11 Tumor Types. J Natl Cancer Inst. 2016;108(11):144.

50. Zhang X, Quan F, Xu J, et al. Combination of multiple tumor-infiltrating immune cells predicts clinical outcome in colon cancer. Clin Immunol. 2020;215:108412.

51. Jiang X, Wang M, Cyrus N, et al. Human keratinocyte carcinomas have distinct differences in their tumor-associated macrophages. Heliyon. 2019;5(8):02273.

52. Yuan ZY, Luo RZ, Peng RJ, et al. High infiltration of tumor-associated macrophages in triple-negative breast cancer is associated with a higher risk of distant metastasis. Onco Targets Ther. 2014;7:1475–1480.

53. Raimondi G, Sumpter TL, Matta BM, et al. Mammalian target of rapamycin inhibition and alloantigen-specific regulatory T cells synergize to promote long-term graft survival in immunocompetent recipients. J Immunol. 2010;184(2):624–636.

54. Fuchs EJ, Matzinger P. B cells turn off virgin but not memory T cells. Science. 1992;258(5085):1156–1159.

55. Parekh VV, Prasad DV, Banerjee PP, et al. B cells activated by lipopolysaccharide, but not by anti-Ig and anti-CD40 antibody, induce anergy in CD8+ T cells: role of TGF-beta 1. J Immunol. 2003;170(12):5897–5911.

56. Ivanova EA, Orekhov AN. Monocyte Activation in Immunopathology: Cellular Test for Development of Diagnostics and Therapy. J Immunol Res. 2016;2016:4789279.

57. Olingy CE, Dinh HQ, Hedrick CC. Monocyte heterogeneity and functions in cancer. J Leukoc Biol. 2019;106(2):309–322.

58. Woods JA, Davis JM. Exercise, monocyte/macrophage function, and cancer. Med Sci Sports Exerc. 1994;26(2):147–156.

59. Lang X, Green MD, Wang W, et al. Radiotherapy and Immunotherapy Promote Tumoral Lipid Oxidation and Ferroptosis via Synergistic Repression of SLC7A11. Cancer Discov. 2019;9(12):1673–1685.

